# Comparison of the ScreenFire and Xpert HPV assays for the detection of human papillomavirus and cervical precancer among women living with HIV in Malawi

**DOI:** 10.1101/2024.02.21.24303142

**Authors:** Chemtai Mungo, Anagha Guliam, Lameck Chinula, Federica Inturrisi, Lizzie Msowoya, Tawonga Mkochi, Siniya Jawadu, Silvia de Sanjosé, Mark Schiffman, Jennifer H. Tang, Jennifer S. Smith

**Author notes:** Corresponding author: Chemtai Mungo.

## Abstract

**Background:** The World Health Organization (WHO) recommends human papillomavirus (HPV) testing for primary cervical cancer screening, including among women living with HIV (WLWH). Low-and-middle-income countries (LMICs) account for 85% of the cervical cancer burden globally, yet have limited access to HPV-based screening, largely due to cost. This study aims to compare the performance of a rapid, isothermal amplification HPV assay (ScreenFire) to that of the Xpert HPV assay for the detection of HPV and cervical precancer among WLWH in Malawi.

**Methods:** We utilized stored self- and provider-collected specimens from a prospective cohort study of WLWH in Malawi from July 2020 to February 2022. Specimens were tested with both Xpert and ScreenFire HPV assays. The overall and within-channel non-hierarchical agreement between ScreenFire and Xpert was determined for both self- and provider-collected specimens. Hierarchical ScreenFire HPV positivity by channel was compared to Xpert for each histological diagnosis - cervical intraepithelial neoplasia grade 2 or worse (CIN2+) compared to <CIN2.

**Results:** 315 matched self- and provider-collected specimens had valid results from both Xpert and ScreenFire testing and were included in analyses. Of these, 245 (78%) had normal pathology, 21 CIN1 (7%), 14 CIN2 (4%), and 35 CIN3 (11%). Among provider-collected specimens, the assays had 80% agreement on overall HPV positivity (unweighted kappa 0.59, 95% 0.50-0.69). ScreenFire was HPV-positive in 90% of self-collected specimens that were HPV-positive on Xpert. Channel agreement between the assays was high for both self- and provider-collected specimens, but slightly lower for HPV18/45. In hierarchical analysis, ScreenFire demonstrated high concordance with Xpert testing for detecting CIN2+ cases in all channels, missing no HPV 16 or HPV 18/45 positive CIN2+ case that was positive on Xpert, in both self- and provider-collected specimens.

**Conclusion:** In this study of stored specimens, the ScreenFire HPV assay performed well in the detection of HPV and CIN2+ among WLWH compared to the Xpert HPV assay. If supported by larger validation studies, ScreenFire could be an affordable alternative point-of-care HPV assay for use in LMICs.

## Introduction

Cervical cancer incidence and mortality data demonstrate a dire global health inequity. Despite cervical cancer being highly preventable, in 2020, low-and middle-income countries (LMICs) accounted for 85% of the estimated 570,000 incident cases and 90% of deaths globally.^1^ The burden of cervical cancer is particularly pronounced in sub-Saharan Africa (SSA), where HIV is endemic.^2^ Women living with HIV (WLWH) have a higher incidence and persistence of high-risk human papillomavirus (HPV), the causative agent of cervical cancer.^2^

The WHO recommends HPV testing as a primary cervical cancer screening method globally, including in LMICs.^3^ Studies on the use of HPV-based screening in LMICs have demonstrated that HPV testing on self-collected specimens is comparable to provider-collected specimens for detection of cervical precancer/cancer,^4,5^ is cost-effective,^6,7^ and highly acceptable across many countries.^8–11,12^

Use of HPV testing for primary screening in LMICs is limited, however, with less than 5 African countries recommending HPV testing as a primary screening method.^13^ This limited use is partly due to the cost and logistical challenges of incorporating HPV testing within existing “screen & treat” programs in LMICs.^14,15^ To increase the feasibility of HPV-based cervical cancer screening in LMICs, affordable, easy-to-use, and point-of-care HPV assays are needed in both facility and community-based screening programs. In addition, the assays ideally need to be able to perform HPV risk stratification through genotyping to inform triaging or management of HPV-positive individuals, based on the carcinogenic risk of each of the high-risk HPV types.^16,17^ This stratification is particularly important in LMICs where the “screen & treat” strategy is used and same-day treatment decisions are often necessary following primary screening to reduce patient loss-to-follow-up.^18^ While several HPV testing technologies with full, extended, or partial genotyping capacity are available, including Aptima® (Hologic Inc, USA), Onclarity (BD Corporation, USA), and Cobas® 4800 (Roche Molecular Diagnostics, Switzerland),^19,20^ the Xpert HPV Assay (Cepheid, Sunnyvale, CA, USA),^21–23^ is one of two WHO-prequalified^24^ HPV tests available for use in field conditions in LMICs. Advantages of the Xpert HPV assay include its ability to detect 13 high-risk HPV types (and HPV66, which is not high risk) in five risk-based channels, which enables risk stratification from a single cartridge in approximately 60 minutes.^25^ The cartridge, which is prefilled with primers and reagents necessary for extraction, amplification, and detection of HPV DNA regions, significantly increases feasibility for use in remote settings without laboratory expertise. Additionally, Xpert® HPV testing is performed on Cepheid GeneXpert platforms, which are widely used for molecular diagnosis of tuberculosis in many LMICs, allowing the integration of both tests on the already available platforms. However, key limitations of the Xpert HPV include the price of the testing (approximately $15/assay,^26^ which is out of reach for most LMICs), the need for a separate PreservCyt medium for specimen preparation prior to testing (which adds logistical challenges and cost), and its inability to simultaneously test a large number of specimens. These limitations highlight a need for alternative assays for use in LMICs.

A promising alternative for rapid, low-cost, and high-volume HPV testing in LMICs is the use of isothermal amplification rather than traditional PCR technology.^27,28^ Isothermal amplification-based HPV assays have been developed by Atila Biosystems (Mountain View, CA, USA) and can potentially be more affordable than most currently available HPV assays.^20^ There are two marketed formats: (i) multiplex detection of 15 high-risk HPV types with separate detection of HPV 16/18 in a single tube (AmpFire HPV assay), and (ii) the Ampfire HPV Genotyping assay with individual genotyping of 15 high-risk HPV types in four tubes.^29^ The assays are Conformite Europeenne (CE) marked^30^ and have been evaluated in analytical^31^ and clinical studies.^20,27,30,32,33,34,35^ These isothermal-based assays have been validated for dry specimen collection,^36^ and allow for storage of dry swabs at room temperature for two weeks,^28^ significantly increasing feasibility for use in LMIC settings.

The existing 15-type isothermal assay (AmpFire HPV) was recently redesigned for public health use as a 13-type assay (ScreenFire) with four channels based on differential cancer risk: (i) HPV 16, (ii) HPV 18/45, (iii) HPV 31/33/35/52/58, (iv) HPV 39/51/56/59/68.^28,37^ This redesigned assay, performed in a single tube, was compared to an AmpliTaq Gold MY09-MY11 PCR-based HPV test on 453 provider-collected samples from Nigeria with very high agreement – the weighted kappa for ScreenFire versus AmpliTaq Gold was 0.90 (95% CI: 0.86-0.93).^28^ Recently, ScreenFire was compared to Linear Array and TypeSeq using 2,076 provider-collected samples from the USA, demonstrating excellent clinical performance, with a CIN3+ sensitivity of 94.7% (95% CI: 92.6-96.4) for ScreenFire, 92.3% (95% CI: 89.7-94.3) for Linear Array and 96.0% (95% CI: 93.9-97.6) for TypeSeq.^37^ Similarly, ScreenFire was compared to Ampfire, Cobas and SeqHPV genotyping using Chinese samples to satisfy Meijer’s Criteria for clinical endpoint validation,^38^ and simulating the VALGENT framework for inter and intra-laboratory validation.^39^ In this study using samples from the Chinese Multi-Site Screening Trial (CHIMUST), ScreenFire demonstrated excellent genotype-specific concordance when evaluated for clinical guidance in hierarchical fashion with both the Cobas 4800 and SeqHPV for <CIN2, CIN2, and >CIN3.^40^

However, there is currently no data available on the performance of the ScreenFire HPV assay compared to Xpert among WLWH in real-world screening specimens. To inform the use of the ScreenFire assay for primary HPV testing in high HIV-burden settings, we compared the performance of ScreenFire with the Xpert assay using stored cervical samples for detection of HPV and cervical precancer among WLWH in Malawi.

## Methods

### Study design, population, and sample collection

We utilized stored self- and provider-collected samples from 315 WLWH in Malawi who participated in a single-arm prospective trial evaluating the feasibility and performance of a same-day HPV-based “screen-triage-treat” algorithm.^41^ The parent study (R21 CA236770) took place between July 2020 and February 2022 at the University of North Carolina (UNC)-Project in Lilongwe, Malawi and enrolled 625 WLWH and 625 HIV-negative women aged 25-50 years. Participant sociodemographic and clinical characteristics were collected at baseline, including HIV status verification, using a rapid (UniGold) and confirmatory (Bio-Rad Geenius) assay. At baseline, HPV testing was done using Xpert from self-collected specimens. Per the parent study protocol, all HPV self-test positive women and 10% of HPV negative women had a pelvic exam to obtain a provider-collected HPV specimen and undergo colposcopy and biopsies (if a cervical lesion was present) or a pap smear (if no lesion was present) for disease status verification. Cervical Pap smears and biopsies were read by two pathologists at the UNC-Project Laboratory and a third pathologist at UNC-Chapel Hill for study quality assurance and adjudication as needed. All patients who were HPV-positive & VIA-positive and eligible for ablative treatment, including those with cervical intraepithelial neoplasia grade 2 or 3 (CIN2/3) on biopsy or high-grade squamous intraepithelial neoplasia (HSIL) on pap smear, were treated with thermal ablation. Participants who were not eligible for ablative treatment and those diagnosed with invasive cancer were referred to the nearby tertiary hospital for care.

HPV sample self-collection was done using a Viba brush (Rovers, The Netherlands), while provider-collection was done using a Cervex broom-shaped brush (Rovers, The Netherlands). HPV testing of dry self-collected specimens using the Xpert HPV was conducted on the same day immediately after sample collection. Residual specimens for HPV-positive samples were aliquoted into cryovials, which were stored at −80°C in ThinPrep at the UNC-Project Malawi laboratory. Provider-collected specimens were sent to the lab, where they were placed into ThinPrep and aliquoted into cryovials. Some of these aliquots were thawed 1-2 years after collection for Xpert testing (when funds became available for such testing) and tested between September 2021 and June 2022. Additional stored aliquots were thawed and tested using the ScreenFire HPV assay between November 2022 and January 2023.

Institutional review board approval for the parent study was obtained from UNC and the National Health Sciences Research Committee in Malawi. All participants consented to specimen storage and future testing.

#### HPV testing by Xpert assay

Specimen testing using the Xpert HPV assay was performed per the manufacturer’s instructions, as previously described.^22^ Briefly, samples were resuspended in 1 ml of PreservCyt, which was added to the Xpert cartridge and placed in the Xpert platform for testing. The Xpert assay tests for 14 high-risk HPV genotypes – HPV 16, 18, 31, 33, 35, 39, 45, 51, 52, 56, 58, 59, 66 and 68. The Xpert results were reported in five channels: (i) HPV 16, (ii) HPV 18/45, (iii) HPV 31/33/52/58/35, (iv) HPV 51/59, and (v) HPV 39/56/66/68.

#### HPV testing by ScreenFire assay

Residual stored specimens were thawed and prepared for ScreenFire testing per the manufacturer’s instructions as previously described.^28^ The ScreenFire HPV assay tests for 13 high-risk HPV types in a single tube, with results grouped based on four risk-based genotype channels: (i) HPV 16, (ii) HPV 18/45, (iii) HPV 31/33/35/52/58, (iv) HPV 39/51/56/59/68. One ml of the preserved samples was centrifuged at maximum speed for 10 minutes to discard the supernatant, and 100 μL of 1X lysis buffer was added to each sample tube. The contents were vortexed and incubated in PCR tubes at 95°C for 20 minutes. After this initial sample preparation, 5 μL of this prepared specimen was mixed with 20 μL of freshly prepared master mix (including reaction mix and primer mix) into a 96-well PCR plate using hand pipetting. Additionally, HPV-positive and negative template controls were included in each 96-well plate. Next, the plates were sealed with an optical compatible film, vortexed gently for mixing the reagents, and centrifuged to bring down all liquid to the bottom of the wells. The plates were then loaded into the Powergene 9600 Plus real-time PCR machine (Atila Biosystems, Mountain View, CA) on the isothermal program mode and run at 1 minute per cycle at 60°C for 60 cycles with fluorescence obtained from CY5 (for HPV 16), ROX (for HPV 18/45), CY5.5 (for HPV 31/33/35/52/58), FAM (for HPV 39/51/56/59/68) and HEX (for Human Beta Globin Gene as Internal Control). A sample was considered positive for a certain genotype group if a signal was detected in the corresponding channel within 60 minutes, regardless of the signal in the HEX channel. If no signal was detected for any of the four HPV channels within 60 minutes, then a signal was required in the HEX channel to be called a valid negative.

#### Analytical Sample

Of 625 WLWH enrolled in the parent study, 295 (47.2%) were HPV-positive on Xpert self-collected specimens, and 292 (98.9%) of those had a paired provider-collected specimen which was stored and tested with both Xpert and ScreenFire assays (Figure 1). Of these, 279 had valid paired self-and provider-collected ScreenFire results after 13 were excluded (2 had invalid Xpert results, 6 specimens were lost, and 5 had invalid ScreenFire results). Per the parent study protocol, every 10^th^ HPV-negative participant on self-collection also had a pelvic exam and a provider-collected HPV specimen, which was also tested on Xpert and stored (N=38). Of 38 Xpert-HPV negatives on self-collected specimens at baseline who had a provider-collected specimen collected and stored, 36 had valid paired results from ScreenFire and Xpert testing after two were excluded for invalid or inconsistent results. In total, 315 (279+36) specimens were included in the analysis. The baseline set of 279 Xpert HPV-positive samples consisted of paired self- and provider-collected specimens, which were tested with both the Xpert and ScreenFire assays. In contrast, for the baseline 36 self-collected Xpert HPV-negative specimens, while the paired self- and provider-collected samples were tested with Xpert, only the provider-collected specimens were available for ScreenFire testing. This difference is because the self-collected specimens for HPV-negatives were not stored after Xpert testing, and therefore, were unavailable for ScreenFire testing. Of the 315 specimens included in this analysis, 245 (78%) had normal pathology, 21 (7%) had CIN1, 14 (4%) had CIN2, and 35 (11%) had CIN3. Sociodemographic and clinical characteristics of the study population are described in supplemental Table 1.

**Figure 1:**
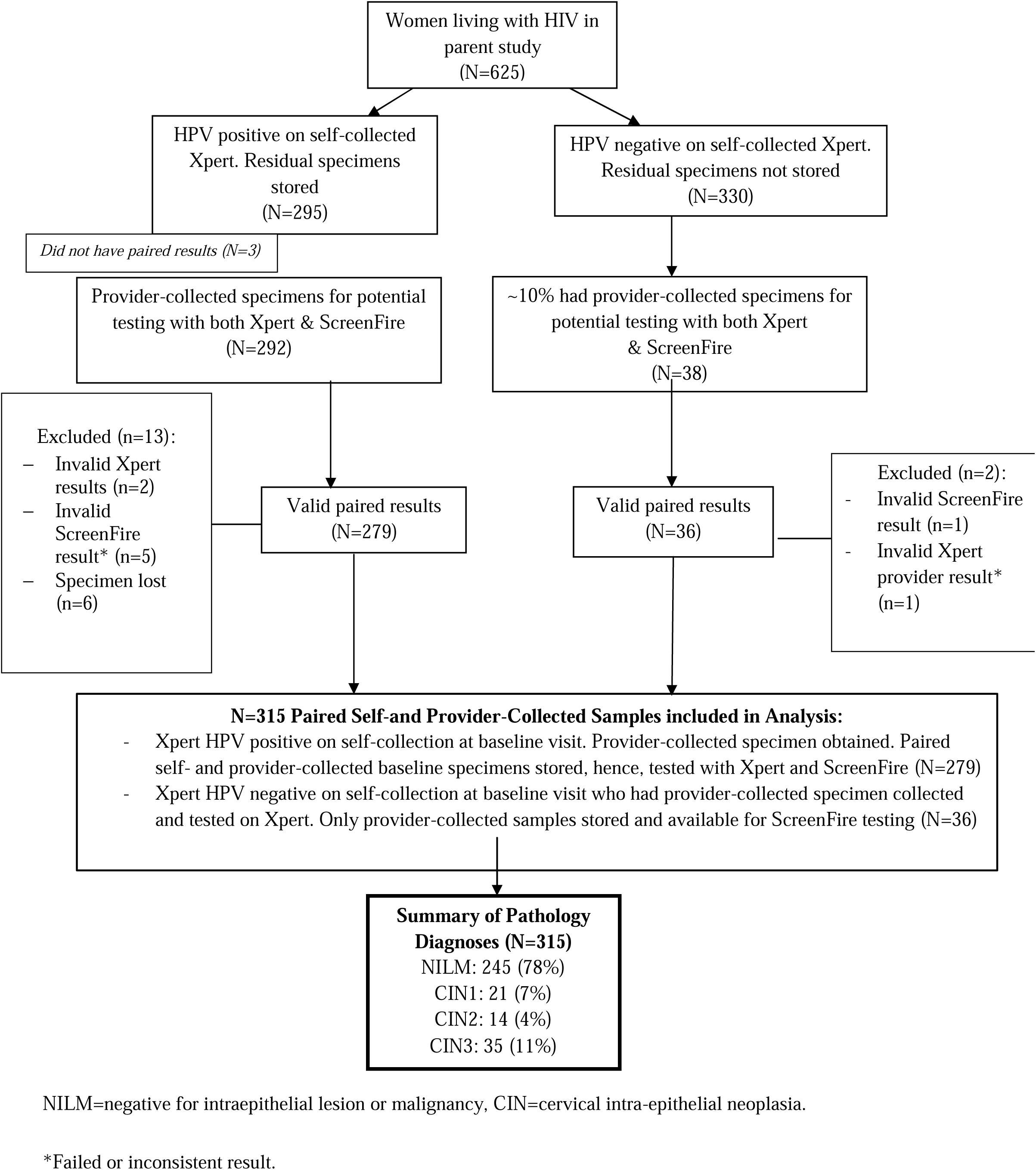
Flowchart of study participants from parent study whose specimens were used in the analysis.

### Statistical Analysis

The overall and within-channel non-hierarchical agreement between Xpert and ScreenFire assays were determined for provider-collected specimens, including percent agreement on positives and corresponding unweighted kappa and 95% confidence intervals (CI). Because only channels 1-3 are similar between Xpert and ScreenFire, percent agreement on positives and kappa were calculated only for these channels. With a similar approach, the overall and within-channel non-hierarchical agreement between Xpert and ScreenFire on paired self-collected specimens were calculated. Because only self-collected specimens were stored from Xpert-positive women and not those with Xpert-negative results, percent agreement and kappa could not be calculated for self-collected specimens.

The two assays were then compared using risk-based hierarchical HPV group types to account for those samples with more than 1 positive channel, considering the different risk of cervical cancer associated with each channel,^42^ using provider- and self-collected specimens. For ScreenFire, the hierarchical risk groups were considered as HPV 16 positive, else positive for HPV 18 or HPV 45 (if HPV16 was not present), else positive for HPV 31/33/35/52/58 (if HPV 16, 18, and 45 were not present), else positive for HPV 39/51/56/59/68, and else negative.^43^ We tabulated hierarchical ScreenFire HPV results and compared them to hierarchical results from the validated Xpert test by histological diagnosis— CIN2 or worse (CIN2+) compared to <CIN2. All analyses were conducted using Stata 17 (StataCorp, LLC).

## Results

Table 1 shows the overall and within-channel agreement between Xpert and ScreenFire HPV tests on the 315 stored provider-collected specimens. Of these 315 specimens, 221 (70%) and 236 (75%) were high-risk HPV-positive on Xpert and ScreenFire, respectively, representing 80% agreement on positives, with an unweighted Kappa of 0.59 (95% CI 0.50-0.69). When comparing agreement within the first three channels, which have identical HPV type groupings, percent agreement was highest (72%) for Channel 3 (HPV 31/33/52/58/35), with an unweighted kappa of 0.73 (95% CI 0.65-0.80), and lowest (60%) for Channel 2 (HPV 18/45), with an unweighted kappa of 0.71 (95% CI 0.60-0.83).

**Table 1:**
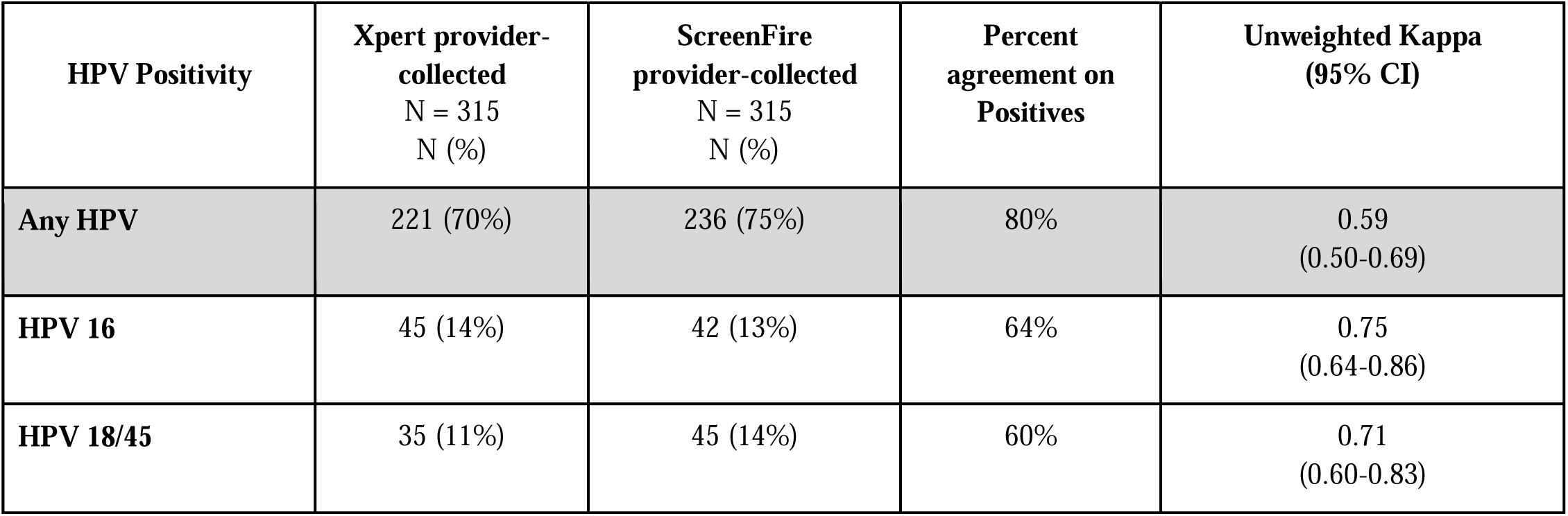
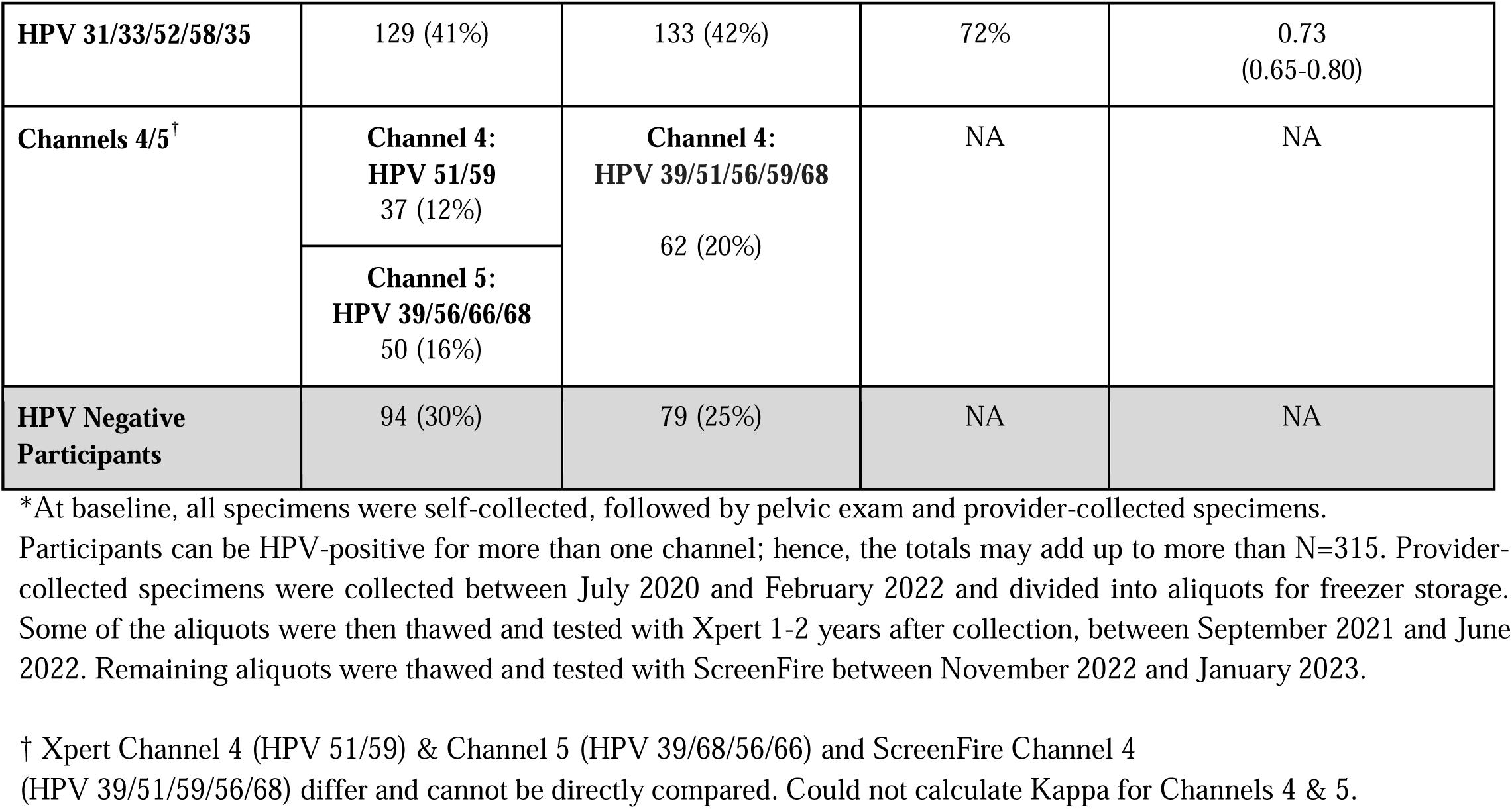
Agreement between Xpert and ScreenFire HPV assays on provider-collected specimens among HIV-positive women in Malawi*.

Table 2 demonstrates the comparison of overall and within-channel agreement between Xpert and ScreenFire on the 279 paired self-collected specimens that were Xpert HPV-positive at baseline. Of these, 252 (90%) were HPV-positive on ScreenFire. Of the 59 (21%) HPV16-positive on Xpert, 49 (18%) were HPV16-positive on ScreenFire. Concordance between the two assays in the HPV 18/45 and HPV 31/33/52/58/35 channels was relatively lower. Of the 49 (18%) HPV 18/45 positive on Xpert, only 38 (14%) were also HPV 18/45 positive on ScreenFire, and of 162 (58%) positive for HPV 31/33/52/58/35 on Xpert, 141 (51%) were positive on ScreenFire. The other channels could not be directly compared as they have different HPV types. A similar comparison was done, stratified by histologic diagnosis (Supplemental Table 2). Because Xpert HPV-negative self-collected specimens at baseline were not stored, percent agreement and kappas could not be calculated for Table 2.

**Table 2:** Agreement between Xpert and ScreenFire HPV assays on self-collected specimens among HIV-positive women in Malawi*.

| **HPV Positivity** | **Xpert**<br>**self-collected HPV positive**<br>**N (column %)**<br>**N=279\*\*** | **ScreenFire**<br>**self-collected result**<br>**N (column %)**<br>**N=279\*\*** |
| --- | --- | --- |
| **Any HPV** | 279 (100%) | 252 (90%) |
| **HPV 16** | 59 (21%) | 49 (18%) |
| **HPV 18/45** | 49 (18%) | 38 (14%) |
| **HPV 31/33/52/58/35** | 162 (58%) | 141 (51%) |
| **Channels 4/5<sup>†</sup>** | **Channel 4: HPV 51/59**<br>52 (19%) | **Channel 4:**<br>**HPV39/51/56/59/68**<br>79 (28%) |
|  | **Channel 5: HPV**<br>**39/56/66/68**<br>65 (23%) |  |
| **HPV Negative Participants** | 0 (0%) | 27 (10%) |

\*All participants were HPV-positive on Xpert by self-collection by study design.
\*\*Participants can be HPV positive for more than one channel; hence, the totals may add up to more than N=279. Self-collected specimens were tested with Xpert in July 2020 immediately after collection and their residual specimen divided into aliquot for freezer storage. The residual specimens were thawed and tested with ScreenFire between November 2022 and January 2023.
<sup>†</sup>Xpert Channel 4 (HPV 51/59) & Channel 5 (HPV 39/68/56/66) and ScreenFire Channel 4 (HPV 39/51/59/56/68) differ and cannot be directly compared.

Tables 3a and 3b show results of ScreenFire hierarchical HPV positivity by channel, stratified by the histologic diagnosis, using Xpert as the reference, for provider- and self-collected specimens, respectively. In these hierarchical analyses, the highest priority cases were CIN2+ with HPV 16. For the hierarchical analyses of provider-collected specimens (Table 3a), among HPV 16-positives by Xpert, ScreenFire detected 12 of 15 (80%) with CIN2+ and 22 of 30 (73.3%) with <CIN2. Here, ScreenFire missed no HPV16 CIN2+ cases identified on Xpert (i.e., 3 HPV16 Xpert positive CIN2+ cases were negative on ScreenFire HPV16 but positive on another ScreenFire channel). Among 30 Xpert HPV16 positive, <CIN2 cases, 22 (73.3%) were also HPV16 positive on ScreenFire, 7 (23.3%) were positive for other channels on ScreenFire, and only 1 (3.35) was HPV-negative on ScreenFire (Table 3a). Among the provider-collected HPV 18/45-positives on Xpert, 7 of 7 (100%) with CIN2+ were also HPV 18/45 positive on ScreenFire, and 21 of 26 (80.8%) with <CIN2 were also HPV 18/45 positive on ScreenFire (Table 3a). For the HPV 31/33/35/52/58 channel, among 20 positives on Xpert with CIN2+, ScreenFire detected 19 (95%) as positive for the same channel, with only 1 (5%) testing HPV-negative on ScreenFire.

**Table 3a:** Hierarchical ScreenFire positivity compared to Xpert for provider-collected specimens among 315 HIV-positive women in Malawi.

| ScreenFire provider-collected<br>hierarchical) | Xpert provider-collected, reference standard (hierarchical) |  |  |  |  |  | total |
| --- | --- | --- | --- | --- | --- | --- | --- |
|  | HPV 16 | else<br>HPV 18/45 | else<br>HPV<br>31/33/35/52/58 | else<br>HPV 51/59 | else<br>HPV 39/56/66/68 | else<br>HPV negative |  |
| **<u>≥CIN2+ \*</u>** |  |  |  |  |  |  |  |
| HPV 16 | 12 | 0 | 0 | 0 | 0 | 1 | 13 |
| else HPV 18/45 | 0 | 7 | 0 | 0 | 0 | 0 | 7 |
| else HPV 31/33/35/52/58 | 3 | 0 | 19 | 0 | 1 | 1 | 24 |
| else HPV 39/51/56/59/68 | 0 | 0 | 0 | 1 | 0 | 0 | 1 |
| else HPV negative | 0 | 0 | 1 | 0 | 2 | 1 | 4 |
| Total | 15 | 7 | 20 | 1 | 3 | 3 | 49 |
| **<u>≤CIN2 †</u>** |  |  |  |  |  |  |  |
| HPV 16 | 22 | 3 | 0 | 1 | 0 | 3 | 29 |
| else HPV 18/45 | 0 | 21 | 1 | 1 | 1 | 11 | 35 |
| else HPV 31/33/35/52/58 | 6 | 1 | 73 | 1 | 3 | 12 | 96 |
| else HPV 39/51/56/59/68 | 1 | 0 | 2 | 12 | 11 | 5 | 31 |
| else HPV negative | 1 | 1 | 5 | 5 | 3 | 60 | 75 |
| Total | 30 | 26 | 81 | 20 | 18 | 91 | 266 |

Note: Highlighted boxes are concordant HPV risk group results between the two assays.
\*Includes 14 cervical intraepithelial neoplasia grade 2 (CIN2), 35 cervical intraepithelial neoplasia grade 3 (CIN3).
†Includes 21 cervical intraepithelial neoplasia grade 1 (CIN1) and 245 with normal pathology

**Table 3b:**
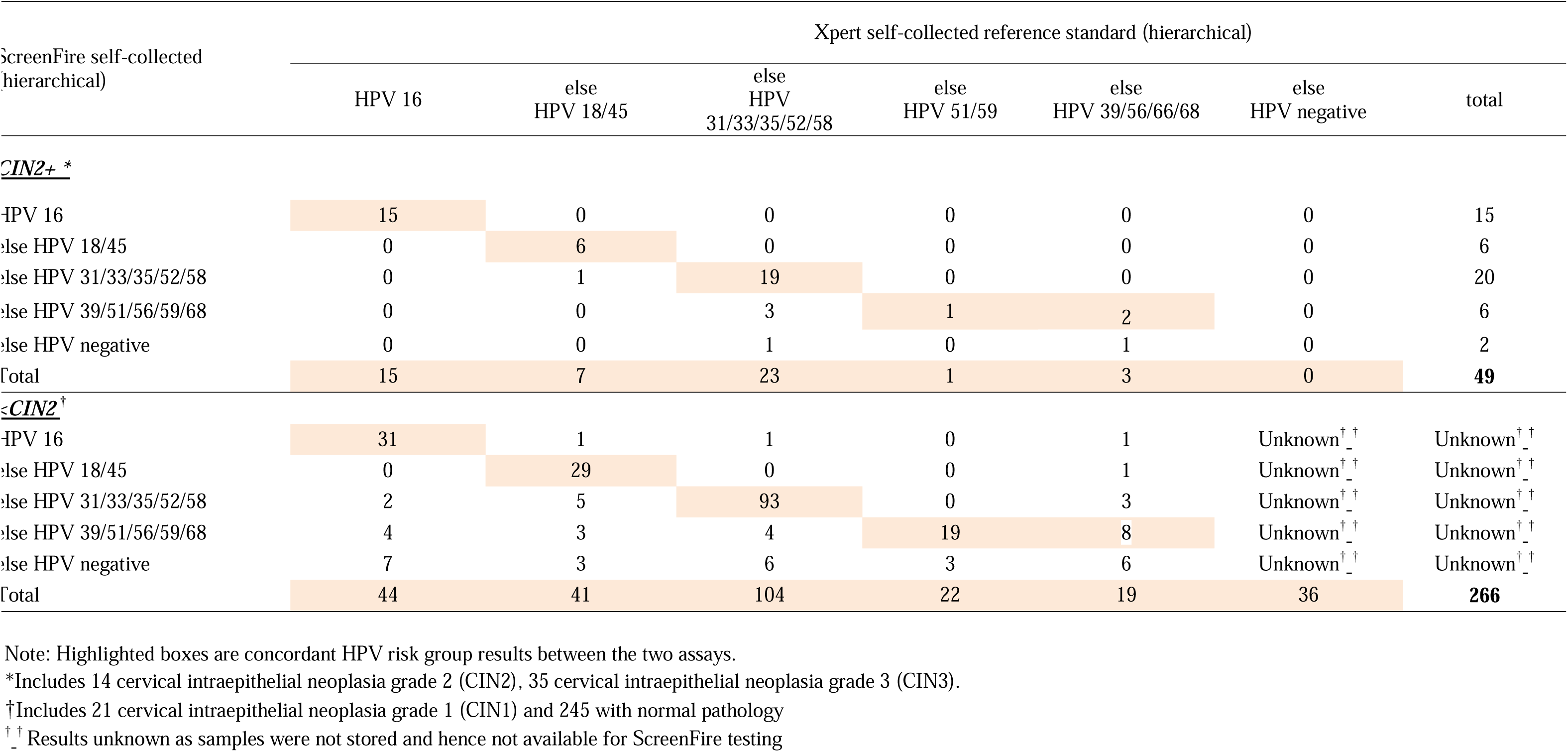
Hierarchical ScreenFire positivity compared to Xpert for self-collected specimens among 315 HIV-positive women in Malawi.

Among the 49 CIN2+ cases in our analysis, all of whom were HPV-positive on self-collected specimens at study entry (Table 3b), 3 (6.1%) were HPV-negative on Xpert provider-collected specimens (Table 3a). Of these 3 CIN2+ missed by Xpert, 1 was also ScreenFire HPV-negative on the self-collected specimen, while 2 were additionally picked up by ScreenFire, with a positive result on Channels HPV 16 and 31/33/35/52/58, respectively. As expected, there was more variation in channel positivity between Xpert and ScreenFire among women with <CIN2, who are known to have lower HPV viral loads than those with CIN2+ for HPV 16 and the HPV 16-related types.^44^

For the hierarchical comparison of ScreenFire and Xpert (reference standard) for self-collected specimens (Table 3b), similarly high levels of agreement are demonstrated. Of 15 CIN2+ cases that were HPV16-positive on Xpert, all 15 (100%) were also HPV16-positive ScreenFire. Among 7 CIN2+ cases that were HPV 18/45-positive on Xpert, 6 (85.7%) were also HPV18/45-positive on ScreenFire, and 1 (14.2%) was positive on ScreenFire HPV 31/33/35/52/58 channel. For 23 CIN2+ cases that were HPV 31/33/35/52/58-positive on Xpert, 19 (82.6%) were positive for the same channel on ScreenFire, 3 (13.0%) were positive on ScreenFire HPV 39/51/56/59/68 channel, and 1 (4.3%) was missed on ScreenFire (tested negative).

In summary, all 49 CIN2+ cases were HPV-positive on Xpert on self-collected samples, while 2 (4.1%) were missed by ScreenFire (Table 3b). Agreement on self-collected specimens between the two assays for the < CIN2 cases was also high for the 3 comparable channels (Table 3b). Among the approximately 10% (N=36) participants who were Xpert HPV-negative on self-collection at study entry and had a provider-collected specimen tested on Xpert and ScreenFire, all 36 were HPV-negative on Xpert provider-collected assay and all had <CIN2 on pathology. None of these Xpert HPV-negative specimens were available for ScreenFire testing. Hence, comparison with ScreenFire could not be done.

Table 3c compares the hierarchical ScreenFire HPV positivity for self-collected specimens to Xpert provider-collected specimens (reference), similarly stratified by histologic diagnosis. Similar to Table 3a, ScreenFire missed no HPV 16 CIN2+ cases that were identified on Xpert. Out of 15 cases of CIN2+ that tested positive for HPV 16 on Xpert, 13 (86.6%) also tested positive for HPV 16 on Screenfire. The remaining two cases tested positive on different ScreenFire channels: one for HPV 18/45 and the other for HPV 39/51/56/59/68. Among 7 Xpert HPV 18/45-positive CIN2+ cases, all were positive on ScreenFire, with 5 (71.4%) also positive on the 18/45 channel.

**Table 3c:**
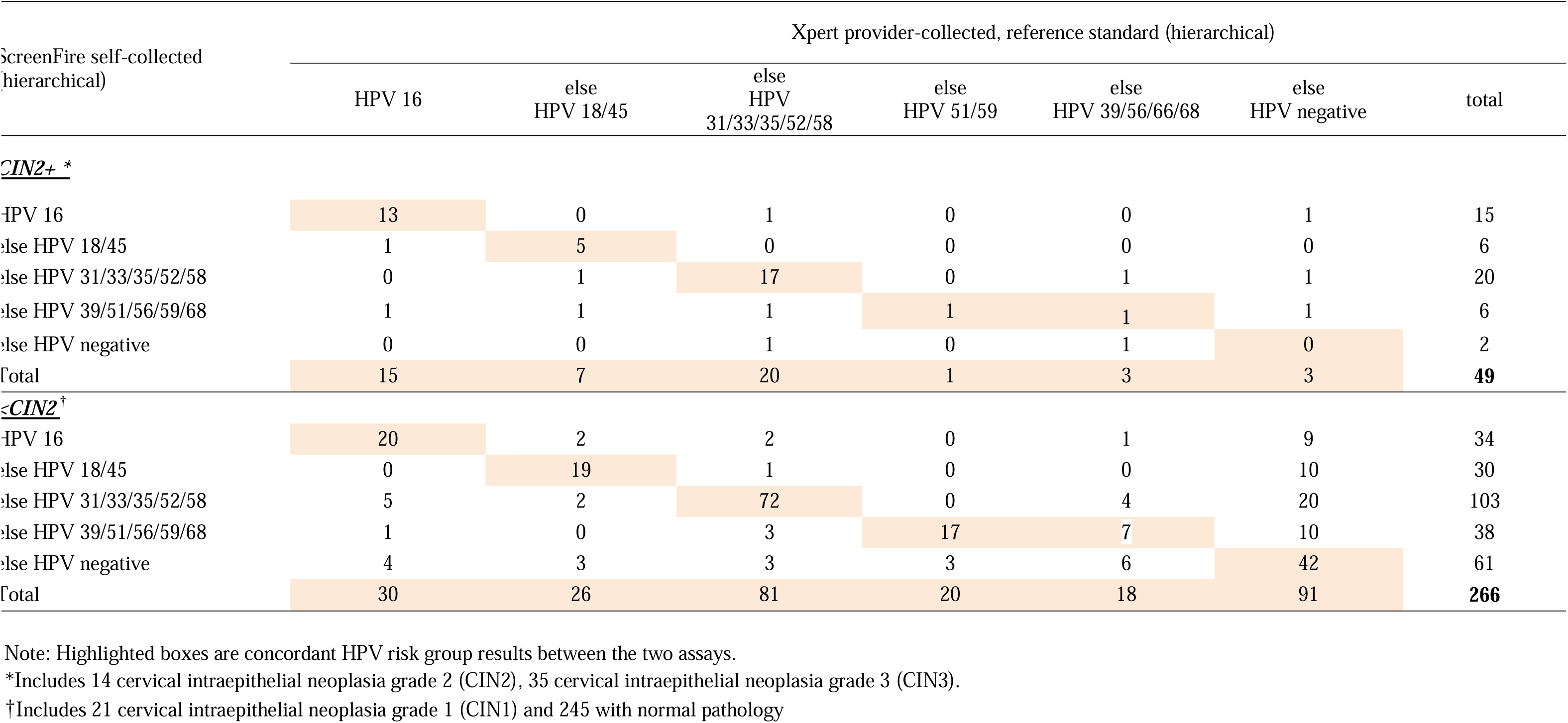
Hierarchical ScreenFire positivity for self-collected compared to Xpert provider-collected specimens among HIV-positive women in Malawi.

In this hierarchical comparison of ScreenFire self-collected to Xpert provider-collected results, there was also high agreement between the two assays for the HPV 31/33/35/53/58 channels. Of the 20 CIN2+ that tested positive for HPV 31/33/35/53/58 on Xpert, 17 (85%) were positive for the same Channel on ScreenFire, and 2 (10%) positive on a different ScreenFire channel (HPV 39/51/56/59/68 and HPV 16). Only 1 (5%) of the CIN2+ cases positive on Xpert on this channel was missed (tested negative) on ScreenFire. Similar to Table 3a, 3 CIN2+ cases were negative on Xpert provider-collected samples, despite being positive on Xpert self-collected specimens. These samples were all positive on ScreenFire on the self-collected specimens.

## Discussion

In this analysis of stored cervical samples of primarily Xpert HPV-positive self-collected specimens among WLWH in Malawi, the ScreenFire HPV assay had good agreement with the WHO-prequalified Xpert HPV assay. ScreenFire demonstrated 80% agreement on HPV positives when compared to Xpert using paired provider-collected specimens. The agreement between the two assays was excellent for the HPV16 channel (unweighted kappa = 0.75) and good for HPV 18/45 (unweighted kappa = 0.71) and HPV 31/33/52/58/35 (unweighted kappa = 0.73) on provider-collected specimens. Among paired self-collected specimens, 90% of samples that were Xpert positive were also ScreenFire positive. Channel-specific concordance was highest for HPV 16 and slightly lower for HPV 18/45. In hierarchical analysis, ScreenFire demonstrated high concordance with Xpert for detecting CIN2+ cases in all channels, missing no HPV 16 or HPV 18/45 CIN2+ case that was positive on Xpert, on both self- and provider-collected specimens. All HPV 16-positive or HPV 18/45-positive CIN2+ cases on Xpert were HPV positive on ScreenFire, although not always for the exact channel. This delineation is important given the importance of HPV 16 and HPV18 in cervical carcinogenesis. Of note, among the 49 CIN2+ cases in our sample cohort, all of which were Xpert HPV-positive on self-collection by study design, 3 were HPV-negative on Xpert provider-collected specimens, of which one was negative on ScreenFire provider-collected specimen. This may be a false negative or possibly sampling error. When comparing the results of ScreenFire self-collected specimens to Xpert provider-collected specimens, all 3 CIN2+ cases that were HPV-negative on Xpert provider-collected specimens were HPV-positive on ScreenFire self-collected specimens.

With the demonstrated comparison, the ScreenFire HPV assay has several advantages that can significantly improve access to primary HPV screening in LMICs where the burden of cervical cancer is highest. ScreenFire can allow testing of up to 96 samples at a time, increasing feasibility for use in community-based campaigns or high-volume clinics at a relatively low-cost (5 USD per test),^27^ which is less than half the price of Xpert HPV tests. Additionally, ScreenFire’s extended genotyping capability, which we demonstrate to have good concordance to Xpert HPV and histology, if supported by larger validation studies, can be utilized for risk stratification in “screen and treat” programs in LMICs.

Compared to some other HPV typing assays available, ScreenFire is relatively simpler to use, does not require DNA extraction, and is compatible with the collection and transportation of dry swabs to the laboratory. These characteristics significantly improve the feasibility of using ScreenFire for extended HPV genotyping in LMIC settings with basic laboratory infrastructure or within community-based and mobile clinics. Limitations of ScreenFire include the need for hand pipetting for sample preparation, which requires basic pipetting skills and individual mixing of amplification reagents, both of which can be subject to contamination under field conditions, reducing accuracy or providing invalid results. Additionally, at ∼6 USD per test, while more affordable than other assays, this price may still be out of reach for many LMICs for use for primary screening.

While the benefits and limitations of the ScreenFire assay have also been discussed in a recent large cross-sectional study, ^37^ our data add two very important aspects specific to LMIC settings. First, this is the first time, to our knowledge, that the novel redesigned ScreenFire HPV assay has been validated on samples from WLWH, who are known to have generally a higher prevalence of multiple HPV infections.^45^ This population is crucial considering the high prevalence of HIV in many LMICs that bear the greatest burden of cervical cancer, and hence could most benefit from high-performing and affordable HPV testing. Second, this is the first time that ScreenFire has been evaluated on self-collected cervicovaginal samples. The use of self-collection is key when implementing primary HPV testing in low-resource settings because of the potential of vastly increasing screening coverage through community or home-based self-collection, when compared to provider-collected specimens, which require a pelvic examination in a clinical setting. Larger studies using ScreenFire on self-collected samples are ongoing, both on WLWH and HIV-negative women.^46^

Among study limitations is the use of frozen samples, some of which had been stored for over two years, which may affect the accuracy of some of our ScreenFire test results. Similarly, because the original study was not designed as a validation study, only Xpert HPV-positive and a random 10% of Xpert HPV-negative specimens on self-collection at baseline were stored and available for ScreenFire testing, possibly biasing our comparison as not all HPV-negative samples were stored for future testing. Lastly, ScreenFire testing in this study was performed at a tertiary-level laboratory in Malawi, with qualified personnel that may not represent field conditions in many LMICs. As such, it is possible that the use of ScreenFire under real-world conditions may yield lower agreement/accuracy compared to our results. To inform this and further guide the use of ScreenFire for HPV testing among WLWH in LMICs, a more extensive study is needed to evaluate test performance and implementation aspects, including the feasibility of testing across different LMICs settings and conditions.

In conclusion, the ScreenFire HPV assay performed well compared to a WHO-prequalified and clinically utilized HPV assay among WLWH in LMIC setting. If supported by larger validation studies, the ScreenFire HPV assay may be a promising option for increasing access to accurate and affordable, risk-based HPV screening and management through genotyping in LMICs.

## Data Availability

Data is provided within the manuscript or supplementary information files. Additional reasonable data requests can be accommodated.

## Acknowledgements

We would like to thank all of the study participants.

---

**Supplemental Table 1:** Demographic and clinical characteristics of included participants.

|  | **HPV-positive on Self-collection by Xpert included in analysis, N=279 n (%)** | **HPV-negative on Self-collection by Xpert included in analysis, N=36 n (%)** |
| --- | --- | --- |
| **Age in years: Median (range)** | 36 (25-49) | 36 (25-47) |
| **Marital Status** |  |  |
| Married | 171 (61.3%) | 25 (69.4%) |
| Divorced/separated/widowed | 101 (36.2%) | 11 (30.6%) |
| Single | 7 (2.5%) | 0 (0%) |
| **Highest Education level attained** |  |  |
| None | 144 (51.6%) | 19 (52.8%) |
| Primary | 100 (35.8%) | 14 (38.9%) |
| Secondary or higher | 35 (12.5%) | 3 (8.3%) |
| **Monthly Income <sup>a</sup>** |  |  |
| < 49,999 MK (< \$47.76) | 189 (67.7%) | 29 (80.6%) |
| 50,000 – 99,999 MK (\$47.76-\$95.53) | 53 (19.0%) | 4 (11.1%) |
| >100,000 MK (> \$95.53) | 37 (13.3%) | 3 (8.3%) |
| **Occupation** |  |  |
| Self-employed<sup>b</sup> | 133 (47.7%) | 20 (55.6%) |
| Salaried employed | 39 (14.0%) | 5 (13.9%) |
| Unemployed /Piece Work | 107 (38.4%) | 11 (30.6%) |
| **Available Household Resources<sup>c</sup>** |  |  |
| Have Electricity | 83 (29.7%) | 8 (22.2%) |
| Have Running water | 94 (33.7%) | 9 (25.0%) |
| **Currently ART** | 262 (93.9%) | 32 (88.9%) |
| **History of smoking, n (%)** | 18 (6.5%) | 2 (5.6%) |
| **Prior cervical cancer screening** | 152 (54.5%) | 22 (61.1%) |
| **Gravidity: Median (IQR)** | 4 (3-5) | 3.5 (3-5) |
| **Number of lifetime sexual partners** |  |  |
| 1 | 28 (10.0%) | 2 (5.6%) |
| 2-5 | 201 (72.0%) | 28 (77.8%) |
| >5 | 46 (16.5%) | 6 (16.7%) |
| Unknown | 4 (1.4%) | 0 (0%) |

<sup>a</sup> MK = Malawian Kwacha. 1 MK = 0.00059 US Dollars as of 16/02/2024 (25,000 MK = 14.85 US Dollars; 100,000 MK = 59.42 US Dollars)
<sup>b</sup> Non-salaried employment e.g. trader, vendor, sex-work, farming
<sup>c</sup> Only those who answered yes are listed.

**Supplemental Table 2:** Agreement between Xpert and ScreenFire on self-collected HPV specimens among 279 HIV-positive women in Malawi*.

| **HPV Positivity** | **Xpert**<br>**self-collected HPV positive**<br>**N (column %)**<br>**N=279\*\*** | **ScreenFire**<br>**self-collected result**<br>**N (column %)**<br>**N=279\*\*** |
| --- | --- | --- |
| **CIN2+** |  |  |
| **Any HPV** | 49 (100%) | 47 (96%) |
| **HPV 16** | 15 (31%) | 15 (31%) |
| **HPV 18/45** | 7 (14%) | 6 (12%) |
| **HPV 31/33/52/58/35** | 34 (69%) | 25 (51%) |
| **Channels 4/5<sup>†</sup>** | **Channel 4: HPV 51/59**<br>8 (16%) | **Channel 4:**<br>**HPV39/51/56/59/68**<br>11 (22%) |
|  | **Channel 5: HPV**<br>**39/56/66/68**<br>12 (24%) |  |
| **HPV Negative Participants** | 0 (0%) | 2 (4%) |
| **<CIN2** |  |  |
| **Any HPV** | 230 (100%) | 205 (89%) |
| **HPV 16** | 44 (19%) | 34 (15%) |
| **HPV 18/45** | 42 (18%) | 32 (14%) |
| **HPV 31/33/52/58/35** | 128 (56%) | 116 (50%) |
| **Channels 4/5<sup>†</sup>** | **Channel 4: HPV 51/59**<br>44 (19%) | **Channel 4:**<br>**HPV39/51/56/59/68**<br>68 (30%) |
|  | **Channel 5: HPV**<br>**39/56/66/68**<br>53 (23%) |  |
| **HPV Negative Participants** | **0 (0%)** | 25 (11%) |

\*Participants were HPV-positive on Xpert by self-collection at study entry.
\*Participants can be HPV-positive for more than one channel; hence, the totals may add up to more than N=279. Self-collected specimens were tested with Xpert in July 2020 immediately after collection and their residual specimen divided into aliquot for freezer storage. The residual specimens were thawed and tested with ScreenFire between November 2022 and January 2023.
†Xpert Channel 4 (HPV 51/59) & Channel 5 (HPV 39/68/56/66) and ScreenFire Channel 4 (HPV 39/51/59/56/68) differ and cannot be directly compared.

